# Navigating a fragmented care system: a qualitative study exploring the combined perspectives of critical care survivors and their clinicians

**DOI:** 10.1101/2025.09.16.25334818

**Authors:** Saira Nazeer, Georgia Mathieson, Zudin Puthucheary, Tim Stephens

## Abstract

Recovery following a critical illness is challenging. Follow-up clinic provision is standard of care, but the effectiveness of services remain unclear, with some data demonstrating limited or potential detrimental effects. While the patients’ perspective has been described previously, no studies combine with clinician experience. We explored the experiences of patients and clinicians to understand modifiable factors in delivering effective post-critical care recovery services.

A single-centre qualitative study was conducted as part of an evaluation of a new care navigator service. Semi-structured interviews were conducted with patients and clinicians, and themes identified.

Sixteen patients and seven clinicians were interviewed. Emergent themes highlighted challenges in coordination between hospital, community and social care teams. Fragmented care was a significant barrier to recovery, resulting in frustration. Clinicians recognised the heterogenous nature of recovery and emphasised the need for patients to receive better support navigating the recovery process.

Our data highlight the importance of integrated follow-up care for critical illness survivors and the need for clinicians to act as navigators within fragmented care systems. Systemic improvements in follow-up care are required, as is the integration of social and healthcare services to ensure that patients receive timely and comprehensive support.

## Introduction

Over 200,000 people are admitted to critical care every year in the United Kingdom (UK) (1). Mortality for patients who suffer a critical illness continues to decrease, as a result of advances in medicine and healthcare, with 70% of patients surviving to leave hospital (2,3). However, for many, this is the start of the recovery journey. Up to 50% of this population survive with new chronic diseases and/or permanent disability, and within a year over half will suffer hospital readmission (4– 9). Physical, psychological and cognitive aspects of recovery, described as ‘post intensive care syndrome’ (PICS), result in greater dependence on health and social care for up to 5 years post discharge (10–12).

Follow-up care for survivors influences recovery by identifying unmet needs and are included in UK national rehabilitation guidelines and provision of care standards (13,14). However, findings from large randomised control trials (RCT) and meta-analyses examining the impact of follow up services have demonstrated little effects on outcomes, with one large RCT finding worsening quality of life (15–17). This highlights the complexities of determining the best service provision for critical care survivors, as they transition across different funding streams and models of care between hospital, community and social care teams. Community teams are often unfamiliar with the critical care recovery process (18). Secondary care teams need to interact and build collaborations effectively with community and social care teams to understand service availability. This places demand on their time acting as patient advocates (19). As the number of critical illness survivors grow, so too does the demand to ensure a smooth transition of care from hospital to community and social care teams.

The post-critical illness recovery journey has been well documented from the patient’s and carers perspective internationally (20–25). Insights on UK services have been limited, focussing on COVID-19 recovery (26–28). However, there are none to our knowledge that look at this in conjunction with the perspectives of treating clinicians, an important insight which may identify mechanisms to improve care and outcomes for this at-risk population.

The aim of this study was to qualitatively explore and describe patient and clinician experiences on recovery post-critical illness, identifying common or divergent themes related to navigating care.

## Methods

### Design

A single-centre qualitative study nested within a feasibility and acceptability evaluation of a pilot follow-up service for critical care survivors. Semi-structured interviews were conducted with patients and clinicians involved in the recovery pathway. Topic guides were developed by the study team (supplementary material file 2), with input from the patient and public involvement group, and included exploration of the post-discharge recovery process. Ethical approval was obtained from the Health and Care Research Ethics Committee (reference number 23/YH/0146, 22.08.2023).

### Sampling and recruitment

Patients were purposively sampled to participate in the interviews, based upon the following criteria: age, sex, ethnicity, primary language spoken and reason for admission. Patient participants were approached once discharged from critical care. Those that agreed to take part in the study were invited for telephone or video interview, scheduled for once discharged from hospital. Clinicians were purposively sampled based on their profession. Written consent was obtained for all participants. Sampling continued until the target 20 patients and 10 staff was achieved or data saturation was reached (29,30).

### Data analysis

Interviews were audio recorded and transcribed verbatim (SN). Transcripts were analysed following reflexive thematic analysis principles (29,30). Analysis commenced after three interviews, with study team members (SN, GM, TS) initially independently coding and then collaboratively developing the code-book to be used for subsequent data analysis (SN, GM), whilst remaining open to identification of new codes. Emergent themes focussing on recovery post critical illness were developed (SN, GM) and then refined by the whole study team in two data meetings (30). The final code-book and core research team positionality is included in the supplementary material (files 1, 3 and 4).

## Results

### Participant characteristics

A total of 16 critical care survivors (including one carer to assist) were interviewed within the first 3 months of being discharged from hospital plus 7 clinicians involved in the critical care follow-up clinic at the study site. Interviews took on average 23 minutes. Participants were recruited between October 2023 and December 2024. Twenty-four patients were approached to take part in the study and provided with patient information sheets. Out of those, two declined due to time constraints, five were lost to follow up and one individual passed away.

### Recovery process themes from interviews

Key themes are summarised below with supporting quotes (Tables 2,3), with a summary infographic (Figure 1)

**Table 1:** Participant characteristics.

| **Survivors (n=16)** | **n(%) or median (IQR)** |
| --- | --- |
| Age at interview (years) |  |
| <40 | 5 (31%) |
| 40-60 | 5 (31%) |
| >60 | 6 (38%) |
| Sex |  |
| Female | 7 (44%) |
| Male | 9 (56%) |
| Length of stay in ICU (days) | 12 (7-15.5) |
| Reason for admission |  |
| Medical | 7 (44%) |
| Surgical | 9 (56%) |
| Ethnicity |  |
| Asian | 2 (13%) |
| Black | 4 (25%) |
| White | 10 (62%) |
| Primary Language Spoken |  |
| English | 12 (75%) |
| Bengali | 1 (6%) |
| Other | 3 (19%) |
| Clinicians (n=7) | n(%) |
| Profession |  |
| Nurse | 2 (29%) |
| Physiotherapist | 1 (13%) |
| Psychologist | 2 (29%) |
| Doctor | 2 (29%) |

**Table 2:** Summary of key patient themes with supporting quotes.

| Patient Themes | Quote # | Supporting quotes |
| --- | --- | --- |
| **Theme 1: Acceptance of slow recovery process as long as some improvement is visible.** | Quote 1 | "You know I just thought it would be you're out. Everything's fine, but it's. It does take time and it's it's it's [sic] a slow recovery...just taking this step by step" (P12) |
|  | Quote 2 | "But it's still a longer process, and I would say, probably everything I do now takes me. It used to take, probably 3 times as long as before. Now probably twice as long, so I guess there is a progress there." (P1) |
|  | Quote 3 | "The curve of recovery, for each injury is slightly different. But luckily, you know, there's always something getting better. So even if I haven't seen, for example, my elbow, I could tell that I am stronger generally. I can start walking. I can start exercising every day you notice that something gets a little bit better. Sometimes it's just overall. You just feel less tired" (P1) |
|  | Quote 4 | "I found it difficult...I was getting out of breath...but I kept walking each day to build it up" (P8) |
| **Theme 2: Getting used to a new level of "what is normal for me"** | Quote 5 | "To some extent my world has shrunk...So you know, you kind of very much limit the range of what you can do. And you know, kind of within that like range, you can function. But it's a limited range comparing to my activities pre accident" (P1) |
|  | Quote 6 | "It's that difference between a realistic recovery and what's going to get better and what's forever going to stay the same. So it's not realistic for me to be in the same position I was in |
|  |  | before because of the spinal cord injury. So some of the times the word recovery was a bit ambiguous in terms of where it was aiming for" (P9) |
|  | Quote 7 | "...which I'm finding it extremely hard to do, because I'm back home right now, and I've got to be a parent again" (P13) |
| **Theme 3: Access to support (including aids/adaptations) can facilitate independence, function and recovery** | Quote 8 | "I am thankful for their {physiotherapists} help they put me on the right course and gave me that little bit of confidence which is you know what you really need." (P5) |
|  | Quote 9 | "I'm writing important things down now on the safety notes." (P13) |
|  | Quote 10 | "My recovery, I mean, has been hampered by me not getting to see, I mean, the specialist I needed to see" (P13) |
| **Theme 4: Acknowledging the impact that critical illness has during admission, discharge and recovery on mental health of patient and family** | Quote 11 | "To be honest, I don't want to go back there. You know. It makes me so anxious just thinking about it." (P1) |
|  | Quote 12 | "It nearly caused the disruption in the family... I don't think he realised what he was taking on...I was left alone in a little room downstairs, and they were mostly being upstairs. I just felt, I just felt really, really terrible. I really thought I was going to die in my own son's house" (P2) |
|  | Quote 13 | "She witnessed the whole thing, and I'm more concerned about her... my daughter's responding quite good to the sessions." (P13) |
|  | Quote 14 | "Although it's a physical thing, it's also a mental thing, you know the recovery is also in your head" (P5) |
| **Theme 5: Frustrations with fragmented care (including patient feeling not in control)** | Quote 15 | "For me, it's been a really exhausting part of recovery, that, that sense that you're on your own, and you've got to fight for a lot of things.....Because there doesn't seem to me to be anyone in overall charge of things. It's quite fragmented. And you don't get the sense that anybody's in overall control, which is me, me, I'm the one in overall control, allegedly" (P9) |
|  | Quote 16 | "You're not always in a fit state to keep track of where it's all gone ...The earlier on the worse you was so for a good half of that I wasn't even fit to realise what was going on" (P7) |
|  | Quote 17 | "...they did MRIs and everything, but they couldn't, they couldn't do any reports on it, because they didn't have |
|  |  | anything to compare with...I don't think the two hospitals were able to talk to each other" (P2) |
|  | Quote 18 | "The doctor did not even ask me why did I come to the surgery? .....not even asking me any question. Or ask me if I have any question for him" (P6) |
|  | Quote 19 | "There's quite there's, there's a lot of opportunities for people to fall through the cracks" (P9) |

**Table 3:** Summary of key clinician themes with supporting quotes.

| **Clinician Themes** | **Quote #** | **Supporting quotes** |
| --- | --- | --- |
| **Theme 1: Heterogenous nature of recovery from critical illness with changing focusses throughout the process and potential for mismanagement.** | Quote 20 | "Definitely critical care. You're very much the acute side of things, basically keeping somebody alive when you're doing the follow up clinic, it's much more rehab, psychological support, referrals that have been for, that have not been done, or that have gone missing, things that you wouldn't really deal with so much in the intensive care setting, I think." (C1) |
|  | Quote 21 | "I imagine that their first thoughts it feels like their first thoughts are the physical but actually the sort of cognitive and psychological seem to make up as much, if not more of the stuff that we talk about [in follow-up clinic]" (C3) |
|  | Quote 22 | "...we pick up the pieces 12 months later and realise it's a bit of a disaster, and they've kind of dropped off from all other services" (C2) |
|  | Quote 23 | "People kind of fall between the cracks haven't been picked up by services. And you know they're struggling much more to get on with things... They're not able to really seek help unless they go into one of our services, and someone picks them up" (C5) |
| **Theme 2: Unfamiliar nature of surviving from a critical illness can lead to unrealistic expectations of recovery and limit a patients' and families understanding of the process** | Quote 24 | "...they don't really understand where they might end up and what they might need to do to get a bit better" (C2) |
|  | Quote 25 | "They're definitely not as aware as then their relatives are more aware of kind of what priority was at that point, and the relative the patient doesn't necessarily realize how sick they were because they couldn't see it" (C1) |
|  | Quote 26 | "Sometimes patients and their families think that I've left intensive care, I've left the hospital, I've had my rehab, and now I'm going home and it's going to be fine. But this is where most of the times the problem starts. And when I'm talking about problems and issues I mean fatigue and nightmares and anxiety and you know I think it's very difficult because it's the first time that they're kind of being alone without any healthcare professionals around them. It's just them and their family and friends. And I think that they don't realise up to that point that they do still have a long road ahead of them and lots of challenges to face" (C4) |
|  | Quote 27 | "Maybe I'll be able to manage this. I just want to get on with my life. They start going back to work. They start, you know, doing all the usual kind of life events and hobbies, etc. And then they start noticing that there are things that they aren't they aren't able to do." (C5) |
| **Theme 3: Impact on, and lack of support for families** | Quote 28 | "Something I have been thinking about more recently is about support for families. And that's kind of related to the fact that often our patients might not have huge amounts of memory of their ICU admission. But their families do, because they were there. They were awake throughout the whole thing. They were the ones who were told that their loved one might not survive or they was, you know, seriously ill that they were, gonna have amputation, or whatever it was." (C6) |
|  | Quote 29 | "I'm fine, you know. I was the patient. I don't really remember or you know. Actually, I'm getting on okay. But it's my, it's my |
|  |  | partner or my son or my nephew that found me unconscious. And they're really struggling" (C5) |
|  | Quote 30 | "We kind of ask families how they're doing, and I think sometimes we do, but then we get a bit stuck, because if they then say actually, I'm not doing so well..... Then we're a bit like Hmm. Okay, what can we do with you now? .... Because I think nationally, that's something that's lacking, anyway, is support." (C6) |

**FIGURE 1:**
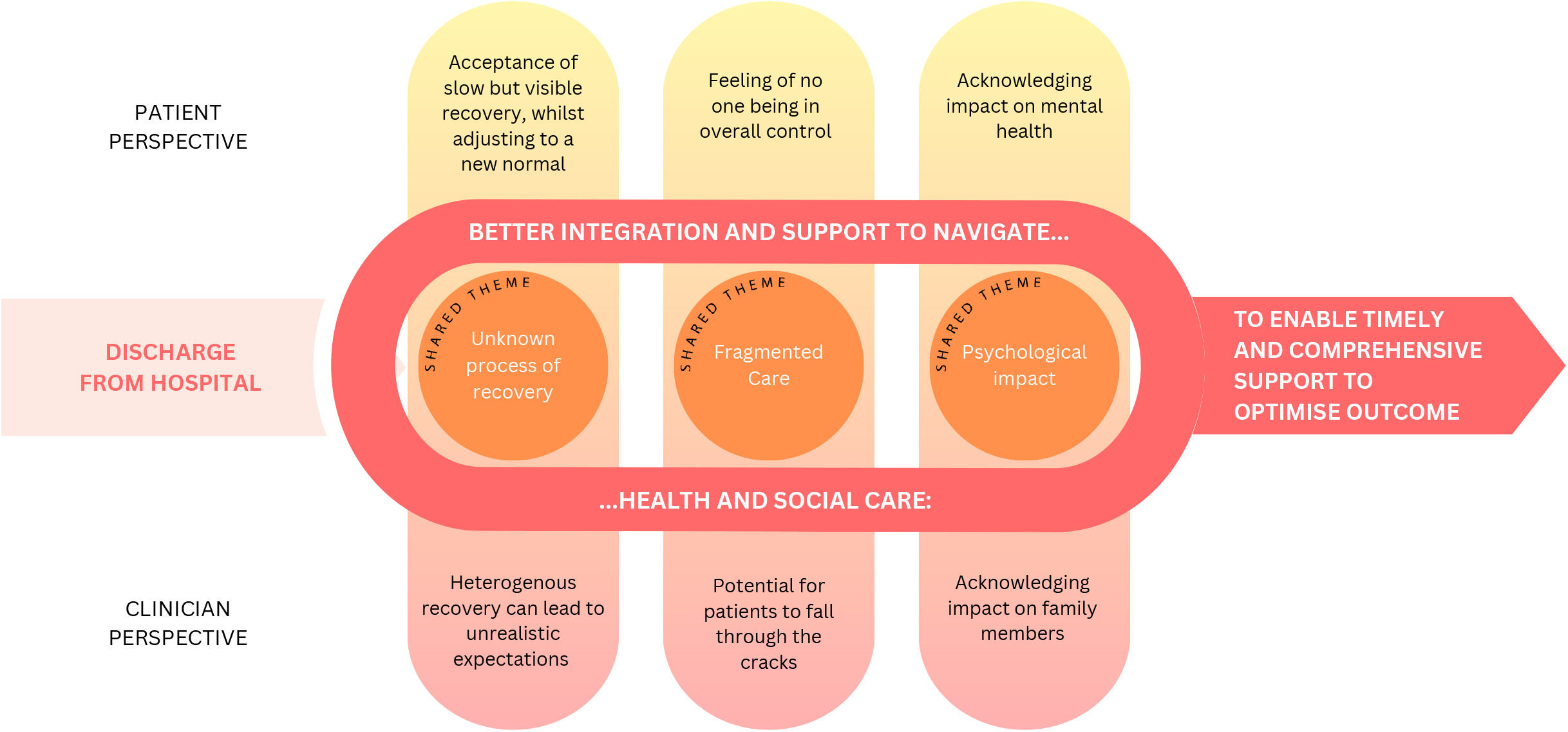
IDENTIFYING KEY CHALLENGES TO PAVE A WAY FORWARD FOR CRITICAL CARE SURVIVORS

#### Theme 1: Acceptance of slow recovery process as long as some improvement is visible

Most participants discussed how their recovery had been a slow process (Quote 1) with one participant stating how this felt particularly true when compared to recovery after non-critical care hospital admissions. Participants described the need to change expectations of how to measure recovery (Quote 2) and how a noticeable improvement in a functional task was extremely encouraging however small (Quote 3). Participants talked about the impact of ongoing symptoms, such as pain and fatigue, and concerns regarding how scars have affected appearance. For the majority, these ongoing symptoms were accepted as part of the process (Quote 4).

#### Theme 2: Getting used to a new level of “what is normal for me”

A recurrent theme was acknowledgment of a “new normal”. When judging their level of recovery, participants were required to decide whether they were measuring progress or comparing how they were before they were admitted to hospital. This differing timepoint for some solicited positive results as they could see that they will get back to their previous level of “normal”. For others, this led to a feeling of acceptance of new limitations (Quote 5) or a change to previous activities they enjoyed, now perceived as too risky due to the possibility of further injuries. Others discussed how there was ambiguity in the word recovery, with some aspects a temporary “normal” and others now their new “normal” (Quote 6).

This adaptation to a “new normal” was anxiety producing for some, leading to reduced social interactions. Some participants attributed this anxiety to the uncertainty of how much recovery will happen. Others discussed the expectation from those around them to return to their previous level of “normal”, whether that be in work or their role in the family (Quote 7).

#### Theme 3: Access to support (including aids/adaptations) can facilitate independence, function and recovery

Many felt grateful for the support from community teams, describing how it led to improved confidence and facilitated their independence (Quote 8). Some felt that this care was tailored to meet their specific needs. Others talked about support and encouragement coming from friends and family.

Some participants talked about adaptations they had come up with to manage symptoms to enable participation in personal care or other daily tasks (Quote 9).

There was an acknowledgement amongst the participants that by not accessing the right support or at the right time, their recovery had been negatively impacted (Quote 10).

#### Theme 4: Acknowledging the impact that critical illness has during admission, discharge and recovery on mental health of the patient and family

The impact of being critically ill on mental health was a common theme. For some, this was acknowledging the mental trauma of the event itself and the admission to hospital (Quote 11). For others this was linked with the discharge process, where they felt they were discharged too early, feeling unsafe initially, and placing pressure on family members acting as informal carers, some who had underestimated this role (Quote 12). The impact on the family member’s mental health and the support they need during this recovery period was also discussed (Quote 13).

There was an acknowledgment that recovering from a critical illness did not just include recovering from physical injuries but also from psychological ones (Quote 14).

#### Theme 5: Frustrations with fragmented care (including patient feeling not in control)

Most patients discussed frustrations with care (or lack of) that they received after they were discharged from hospital. One participant shared how mentally fatiguing this was to become the person with overall control of a complex set of new care needs (Quote 15). This was particularly difficult in the context of managing multiple follow-up appointments, made harder by the fact that this responsibility was running concurrently with recovery from being critically unwell (Quote 16). Others discussed the frustration of dealing with fragmented care providers who seemingly didn’t communicate (Quote 17), with some feeling that there was no communication of information from their critical care stay, leading to disappointing interactions with their GP who from their perspective hadn’t demonstrated an awareness of the recovery process (Quote 18). Worries were expressed that this fragmented care could lead to not getting the support services needed and therefore impacting on recovery (Quote 19).

### Clinician themes

Most of the clinicians that were interviewed had a unique standpoint in that they interacted with this patient population at two timepoints in their recovery journey: at the beginning on the critical care unit, and then in the critical care follow up clinic. The time at which patients are seen in this clinic varies from as early as 3 months post discharge from critical care to over a year post discharge.

#### Theme 1: Heterogenous nature of recovery from critical illness and changing focus throughout the process leading to risks of potential for mismanagement

Clinicians expressed that the heterogenous recovery process meant that it was difficult to predict the trajectory and have realistic conversations about recovery. They described how there was a difference in focus of recovery on the intensive care unit compared to in follow up clinic. This was described as an initial focus on the physical aspects of recovery to then the more psychological aspects once they have been discharged into the community (Quote 20). They discussed this shift not only from the point of view of managing patients but also for the patient themselves (Quote 21).

This heterogenous nature of recovery places patients at risk of not being identified by the right services to address their needs. The need to prevent mismanagement and patients “falling through the cracks” was discussed by clinicians as their role in the follow up clinic (Quotes 22, 23).

#### Theme 2: Unfamiliar nature of surviving from a critical illness can lead to patients’ and families having limited understanding and unrealistic expectations of recovery process

Surviving a critical illness is an unfamiliar process to patients. Clinicians described how patients often do not have a clear understanding of recovery trajectory and potential for recovery of function nor how long it will take when discharged from critical care and hospital (Quote 24). Clinicians shared that for many patients this may be because they have little memory of their stay on critical care, and that discussions about expected recovery were primarily had with their family (Quote 25). They also reflected on whether these conversations were even being had on critical care whilst trajectory was still unclear, and then it is unknown what happens once they have been discharged to the ward.

Patients sometimes had unrealistic expectations about recovery, with a sense that once they are home they will feel better (Quote 26). Clinicians described how patients may even start to reintegrate back into their previous societal roles, including work, but then acknowledge that they still need support to continue to recover (Quote 27).

#### Theme 3: Impact on, and lack of support for families

The final clinician theme that emerged was recognition of the impact having a relative in critical care has on the family (Quote 28). The patient themselves may alert their clinician to this (Quote 29) or it is identified in follow up clinic. Clinicians commented on the fact that although they may be identifying support needs for families in practice, nationally there is a lack of support available for relatives of critically unwell individuals (Quote 30).

## Discussion

We set out to explore and describe patient and clinician experiences and views on recovery following a critical illness, identifying common or divergent themes related to navigating the recovery process. Three main shared themes emerged. Firstly, the effect of recovery being an unknown process: for the patients they felt that a slow recovery was acceptable as long as some improvement was visible and that this recovery included a process of getting used to a “new normal”. Clinicians expressed that the heterogenous recovery process meant that it was difficult to predict the trajectory and therefore have realistic conversations. This risked these conversations not happening, contributing to uncertainty once the patient leaves critical care. The second shared theme was the impact of fragmented care: patients expressed their frustration with this, feeling that no-one was in overall control and the clinicians acknowledged the potential for patients to fall through the cracks. The final shared theme was the importance of acknowledging the mental health impacts on both patients and their families. Patients discussed the importance of access to support (including aids/adaptations) as a facilitator of independence, function and recovery. Clinicians discussed a shift in focus from physical disabilities to psychological disabilities later in the recovery process.

### Implications for clinical practice

The heterogeneous nature of the patient population, their social circumstances, and their medical conditions result in a variation in response to surviving a critical illness. This in turn leads to heterogenous symptoms that survivors face, exacerbated by fragmented care systems across secondary, primary and community care. The result is that survivors struggle to receive the support they need, when they need it. Better integration and support to navigate health and social care is needed to ensure timely and comprehensive support. Our data offer three clear insights into changes into clinical practise that can improve patient care. Firstly, that critical care follow-up clinicians play an important role not only as care deliverers, but also as care navigators, a key step in understanding how to improve the effectiveness of the recovery process for this patient group. Secondly, discussions about the future of recovery need to be managed earlier in the process, providing an opportunity for expectation management with patients and family. Lastly, in keeping with the extensive published literature, this population is at risk of developing new physical, psychological and cognitive impairments requiring early identification via a standardised tool and a mechanism to operationalise this at a systems level (7,9,10,31).

### Implications for future research

Future research should look to address the above three outlined implications for clinical practice by investigating the role of clinicians as care navigators, determining an appropriate tool to identify needs and then understanding how best to operationalise this. One factor highlighted in our data that needs to be considered is the response shift that occurs during recovery, described by the patient participants as an acceptance of the “new normal”. A response shift is a phenomena where a change or an adjustment in the way a patient perceives their quality of life and health status is observed (32). It can be reflected in patient related outcome measures (PROMS) and thus needs to be taken into consideration when analysing data from PROMS over time, especially if using the results to influence decision making and policy (33). Although this response shift has been observed in the critical care survivorship literature, we are yet to understand how it may be influenced so that clinicians can support patients to improve their perceived quality of life (21,34). This in turn may improve discussions about recovery and expectation management, improving communication between clinicians and patients/family. Future research needs to explore the underlying mechanisms of this response shift observed during recovery, so that those supporting critical care survivors (e.g., clinicians, family members, policy makers, researchers) can understand what can be done to influence perceptions of health.

One criticism of the current literature is that research is hospital-centric, suggesting that service providers should look beyond this at the complex matrix of support services and the way in which healthcare and social care interacts (35). As clinical systems and reimbursement mechanisms differ internationally, there are limits to how much we can extrapolate learnings (36,37). Research into this field therefore needs to be country and situation specific, with future research focussing on the interaction between health and social care and how that impacts on the recovery experience. This will help clinicians and critical care survivors navigate these systems enabling timely access to care. Without this the responsibility will continue to sit with patients and family members to navigate across disjointed systems, leading to unrealistic expectations and frustrations with fragmented care (38).

### Strengths and limitations

There is a wealth of published data from patients on their experience of the recovery process post critical illness (20–28). To our knowledge this is the first study combining the clinician and patient perspectives and examining key commonalities and differences. These data build on a body of work exploring perspectives to assist development of systems which enable clinicians to support patients in navigating health and social care teams. The main limitations of this study are that it is single centre, leading to a small sample size of clinicians and that it lacks the voice of the family/carer, an important insight to comprehensively understand this recovery process (39). Multi-centre research is needed to enhance our understanding of how this process can be improved at a multi-system level, ensuring funding to include the carer’s voice alongside patient and clinicians.

### Conclusion

Our data highlight the variability in recovery experiences, the need for early discussions about recovery expectations, and the frustrations and potential risks with a fragmented care system. These findings indicate a pressing need for systemic improvements in follow-up care and the integration of social and healthcare services to ensure that patients receive timely and comprehensive support.

## Data Availability

Patient and Clinician codebooks are included in the supplementary material. Complete data are available upon reasonable request to the authors.

## Acknowledgements

We would like to thank the patients and staff who were interviewed for this study for their time and sharing their experience with the researchers. We would also like to thank Joe Russell for assisting with the development of the infographic.

## Authors’ contributions

SN, TS and ZP conceived and designed the study.

SN and GM recruited the participants for the study.

SN and TS were involved in the collection of data.

SN, GM and TS performed the analyses.

All authors read and approved the manuscript.

## Data Availability Statement

The data that supports the findings of this study are available, in an anonymised manner, from the corresponding author upon reasonable request.

