## Supplementary Material for "Navigating a fragmented care system: a qualitative study exploring the combined perspectives of critical care survivors and their clinicians"

**Contents:**

**File 1: Core Research Team Positionality**

**File 2: Topic Guides for Semi-Structured Interviews**

**File 3: Patient Codebook**

**File 4: Clinician Codebook**

**File 5: COREQ Checklist**

**File 1: Core Research Team Positionality**

TS is a clinical lecturer, qualitative researcher and critical care nurse. Prior to this study he had no significant experience in, or views about, post-intensive care syndrome or the post hospital recovery process for this patent group.SN is a research physiotherapist who has worked in critical care, in the critical care follow up clinic and, in this study, as a care navigator for critical illness survivors. Until this last role she had little experience of, or strong views regarding, post hospital recovery. SN conducted all patient and clinician interviews (the first 3 peer reviewed by TS). GM is a junior doctor working on the adult critical care unit, who was also involved in recruitment for this study. Prior to this, she had little experience of the post hospital recovery process for this patient group. ZP is a critical care consultant and an expert in post-intensive care syndrome. He did not take part in theming.

**File 2: Topic Guides for Semi-Structured Interviews**

Semi-structured interviews were run as part of a wider service evaluation, and thus most of the questions focussed on feasibility and functionality of the service. The initial line of questioning however prompted a more narrative discussion about the recovery process. It is from this that themes were developed.

**Patients:**

1	How has your recovery been since we last spoke? Or 'Tell me about your recovery so far'

*(Allow participant to describe recovery in a narrative way – subsequent questions will be asked to clarify key issues after this initial narrative summary)*

2	PROSPER feasibility and ease of use

2.1 Have you completed the ePRO questionnaire in the last 4-6 weeks?
2.2 Overall, How was completing the ePRO questionnaire??
2.3 Did you find the online system easy to use? (NB: not for patients using telephone version)
2.4 Did you understand all the questions? If no, please elaborate as best you can
2.5 Roughly how long did it take you to complete the ePRO? Did you find the time taken to do this acceptable to you?
2.6 Is there anything you would suggest we could do to make completing the questionnaire easier for you?
2.7 Did you ever have any help from other people to complete the online questionnaires?
2.8 Do you think there are things missed out in the questionnaires, which could make a difference in terms of supporting your recovery? If so, please elaborate
2.9 Do you think the translated questionnaires were easy to understand and interpret?

3 ePRO functionality questionnaire
  3.1 Did you report any complications (symptoms) using the ePRO questionnaire? (NB If no, end questions here)
  3.2 Did you get any help or advice from our team after you completed the ePRO questionnaire?
  3.3 If yes, please briefly describe if you have had any telephone contact with any health care services in the last few weeks? If yes, who were you in contact with, for what reason and what was the outcome? Was this contact as a result of you completing the ePRO?
  3.4 Did you see any health care professional in person in the last week? If yes, who were you in contact with, for what reason and what was the outcome? Was this contact as a result of you completing the ePRO?
  3.5 Are you glad that you took part in the service?
  3.6 Do you think that completing the ePRO has made a difference to your recovery so far?

**Clinicians:**

1.1. What is your role?
1.2. What stage of their recovery process do you see patients in? If multiple stages, do you think there’s a difference in perspectives at these different stages regarding recovery?
1.3. What part of the patient pathway you work on was affected by the ePRO project?
1.4. Overall, did you find the ePRO project helpful to your role? Please elaborate
1.5. Overall, did you find the ePRO project helpful for patients you cared for?
1.6. From your perspective, were there any challenges created by the ePRO project?

1.6.	Do you think there are things missed out in the questionnaires, which could make a difference

in terms of supporting your patient’s recovery? If so, please elaborate

1.7.	Do you have any suggestions for improving the utility of the ePRO system?

**File 3: Patient Codebook**

| **CODE** | Cited in | **SUPPORTING QUOTES** |
| --- | --- | --- |
| **RECOVERY PROCESS** | | |
| **Theme 1: Acceptance of slow recovery process as long as some improvement is visible.** | | |
| 1.1 having realistic expectations and accepting a slow and steady but visible recovery | P1, P1, P14, P2, P3, P12 | “it is sort of a weekly in other just gradually, gradually, gradually getting back to normal” (P14)  “I wish it was all going faster, but you know you can't really rush his things. It was a pretty horrific accident.” (P1)  “But it's still a longer process, and I would say, probably everything I do now takes me. It used to take, probably 3 times as long as before. Now probably twice as long, so I guess there is a progress there.” (P1)  “slow I would say...but then that was the nature of the accident wasn’t it” (P2)  “know I just thought it would be. You're out. Everything's fine, but it's. It does take time And it's it's it's (sic) a slow recovery…just taking this step by step” (P12) |
| 1.2 Daily improvements visible with functional tasks and previous activities becoming easier | P1, P1, P14, P8, P3, P13, P12, P5, P5 | “So obviously comparing to the way I was just after the accident, you know, completely immobile, not being even able to scratch my nose or even press a buzzer than obviously this is great. I am. I would say. Almost independent to a large extent, but of course, comparing to pre-accident, I was still a long way to go” (P1)  “I do a little bit more and a little bit more after my park run time has come down to 31 1/2 minutes.” (P14)  “But at the same time, as I said, I do see an improvement. Almost, you know, daily, but something I can do. Maybe my elbow I still can’t raise. But I can lift my leg. I can bend my knee, you know, getting into the car. If I go with friends somewhere, it's not a problem. I can walk without a crutch, I can negotiate the stairs, up and down reciprocally, you know, which is, which is a great thing” (P1)  “So obviously comparing to the way I was just after the accident, you know, completely immobile, not being even able to scratch my nose or even press a buzzer than obviously this is great. I am. I would say. Almost independent to a large extent, but of course, comparing to pre-accident, I was still a long way to go” (P1)  “I’ve kept walking a lot each day and gradually built it up” (P8)  “generally day to day she’s up walking around, she is seeing friends and things like that as well now” (P3)  “now I can get in a bath, so I am progressing, yeah, not at the pace I would like” (P13)  “I feel better. But you know it's only because I can sort of get out of bed and. Sort of go for walk. And just sort of. You know, get active” (P12)  “I had one crutch when I seen him and he said to me now I can do without the crutches. So I can do now, I can start driving again so you know once I start driving again I can do better with my shopping and things like that” (P5)  “know I do the exercises myself each day I walk each day, I walk further each day” (P5) |
| 1.3 Multiple injuries/ ongoing symptoms, so different things are recovering at different speeds | P1, P1, P2 | “The curve of recovery, for each injury is slightly different. But luckily, you know, there's always something getting better. So even if I haven't seen, for example, my elbow, I could tell that I am stronger generally. I can start walking. I can start exercising every day. You notice that something gets a little bit better. Sometimes it's just overall. You just Feel less tired” (P1)  “I wish it was all going faster, but you know you can't really rush his things. It was a pretty horrific accident.” (P1) |
| 1.4 Acceptance of ongoing symptoms during recovery | P1, P15, P7, P8, P13, P5 | “I think to some extent you learn to live with it “ (P1)  “Okay, everything's exhausting. But yes, nice of being home… people think its all better when you do get home but its just like the next step really” (P7)  “I found it difficult...I was getting out of breath...but I kept walking each day to build it up” (P8)  “I still can’t go upstairs…But it would be good to get upstairs as then I can get up and sit in the shower and have a proper wash down with the help of my wife so yeah so apart from that I’m doing really well.” (P5)  “It’s a work in progress as well isn’t it” (P5) |
| 1.5 Slower progress with recovery than expected when compared to recovery post admission from chronic condition | P6 | “It's a bit slower than what I expect. Because…When I have a crisis. And I come home after an admission. By now. I would have been doing some things by myself. but this time around I still depend a lot on people.” (P6) |
| 1.6 Pain/ fatigue continues to limit daily activities | P3, P13 | “She's still really tired sleeping a lot. She you have to really encourage her to get up, and you know you can't just sleep. Let's do this. Let's do that. She's very as not cooperate or cooperating, but she just says I can't. It's too painful. Her leg is her leg more than her neck. She's complaining, and her leg hurts more than her neck.” (P3)  “The main concern for me is, you know, when I'm walking or when I get up, I just got pain in my right hip” (P13) |
| 1.7 Ongoing concerns about cosmetic appearance | P3 | “You know she's got a very you know about her face and her scars. She's quite aware of them….I look ugly” (P3) |
| **Theme 2: Getting used to a new level of ‘ what is normal for me’** | | |
| 2.1 Learning to adapt to a new disability but it’s a slow learning process | P9, P12 | “Trying to find things that work. It doesn't necessarily, it's not fixed, but I'm still trying to find ways to work around it.” (P9)  “It's it does require new outlook on what I've got to do” (P12) |
| 2.2 Judging recovery depends on what ‘now’ is being compared to, whilst in hospital or before hospital | P1, P1, P9 | “You kind of very much limit the range of what you can do. And you know, kind of within that like range, you can function. But it's a limited range comparing to my activities pre accident.” (P1)  “But obviously I do see there's still a really long way for me to go. You know I would love to just get out of bed and just go for a jog, you know. Not that I like doing it. But now it seems like a luxury just to be able to do it, and just do other stuff. Just lift heavier things that I was quite strong before the accident. But you know I just try not to look back. I just tried to see. You know, I had a really low base after the accident. And now I just need to kind of, you know, negotiate it. And to an extent I’m improving.” (P1)  “It's that difference between a realistic recovery and what's going to get better and what's forever going to stay the same. So it's not realistic for me to be in the same position I was in before because of the spinal cord injury. So some of the times the word recovery was a bit ambiguous in terms of where it was aiming for” (P9) |
| 2.3 Acknowledgement that things may never be quite the same again – will there need to be some level of acceptance of new limitations | P1, P1, P14, P7, P13 | “Obviously, when the point comes, when I reach a plateau, then depending how high that is or how low, then you know it may be a different story, whether I need to acknowledge and resign my life as always will be limited to some extent.” (P1)  "It is sort of a weekly in other just gradually, gradually, gradually getting back to normal, except hearing, of course, but I've got enough hearing to hear you. So that's not it, not the end of the world at all, not complaining about it" (P14)  “But obviously my elbow is, you know, to what extent I can, if I can just bend it enough and be able to use it so I can touch my face, brush my teeth, eat with my right hand, you know. That's the worry.” (P1)  “Everything’s hard work, but the things that you’re not used to being hard work” (P7)  “But, you know, sometimes you've got to sit down and say, well, it is what it is” (P13) |
| 2.4 Change to doing certain activities now perceived as risky to reduce the risk of another injury | P1, P5 | “I may not you know end up skiing again, but to be honest I don't want to have another injury or something. I think I'm getting more worried now about having an issue, so I'm sure my risk appetite will be much lower after this.” (P1)  “I still can’t go upstairs. So although I am walking without crutches, the physio didn’t want me to go up the stairs without until there’s the other arm rail“(P5) |
| 2.5 Acknowledging new reduced baseline but not attributing this to critical illness/ ICU stay | P8 | “I’m fine now back to my own self, but a tiny bit slower than I was walking…I don’t know where that has come from” (P8) |
| 2.6 Not knowing how MUCH recovery will occur can be anxiety producing | P1, P1, P1, P9, P13, P12, P12 | “You don't know the end line. You don't know how far you can go, you know if somebody told me it's just a question of time and patience, you'll be absolutely fine in a years’ time, I can wait, you know. But you kind of…the worry, and I guess the anxiety is how far can you recover.” (P1)  “But obviously my elbow is, you know, to what extent I can, if I can just bend it enough and be able to use it so I can touch my face, brush my teeth, eat with my right hand, you know. That's the worry.” (P1)  “I guess nobody really knows, because, you know, the exact science all depends on the individual, depends how well your body responds, how you know, obviously, physiotherapy goes and how much you put effort in it.” (P1)  “It's that difference between a realistic recovery and what's going to get better and what's forever going to stay the same. So it's not realistic for me to be in the same position I was in before because of the spinal cord injury. So some of the times the word recovery was a bit ambiguous in terms of where it was aiming for” (P9)  “Yeah, she's referred me for MRI because I was coming down the steps this morning, and I've got a twinge in my lower back. So once I get that, I can know, I mean, you know, if, if everything's right, then I can go into, you know, training my thighs and my legs until I find out what's wrong with my left shoulder. So it’s a step in, in the right direction, so to speak, yeah, give me peace of mind, which I don't have right now.” (P13)  “Sort of knowing that they're on their way, and they could come at any time, is worrying as well” (P12) |
| 2.7 Accepting that during recovery 'my world has shrunk' | P1 | “To some extent my world has shrunk...So you know, you kind of very much limit the range of what you can do. And you know, kind of within that like range, you can function. But it's a limited range comparing to my activities pre accident” (P1) |
| 2.8 Is a realistic recovery getting back to work? Goal to get back to work. | P9, P8 | “I should have been back at work last week, but they haven't had the report from occupational health yet …I'd love to get back to it. …The GP recommended 62 days phased return.” (P8) |
| 2.9 Reduced social interactions | P3, P13 | “Once my kids go to school in the morning, I don't want anybody you know, with me. I don't feel comfortable with it” (P13)  “Sometimes my phone rings, I don't feel like answering to be quite honest. You know, just tired of being you know, what can I say? I mean, people want to speak to me. Sometimes I just don't feel in the shape of mind to talk“ (P13) |
| 2.10 Demand/expectation to return to previous role/stature in family/community | P13, P13 | “Which I'm finding it extremely hard to do, because I'm back home right now, and I've got to be a parent again” (P13)  “I mean, for the sake of the kids, really, so I've got to do whatever I got to do” (P13) |
| **Theme 3: Access to support (including aids/adaptations) can facilitate independence, function and recovery** | | |
| 3.1 Using adaptations and/or devices can mean you can achieve personal care tasks activities of daily living independently | P1, P13, P12 | “So I've been doing it myself, but I still can’t do it properly, because of my arms. So my right arm doesn't bend. My elbow is kind of at a 90 degree, which means I can't really use my right arm to help wash myself, wash my hair. So you kind of you know you learn to live with it and use certain props” (P1)  “I'm writing important things down now on the safety notes. So, you know, I mean, if it gets, if it gets intolerable and I can't function, then of course, I will accept help” (P13)  “It's just watching what I'm eating, and just make sure I'm eating regularly and try not to overeat, but also try not to undereat. And just sort of trying to pace everything, you know, just when I go out and make sure I've got sort of energy bars or snacks, or an energy drink something that I can take with me to sort of so I’ve got food on the go. You know, if I, if I need to go up to Canary Wharf, or something like that” (P12) |
| 3.2 Seeing the value in rehabilitation activities even though it’s painful /unpleasant | P1 | “The problem is that you have to kind of brace yourself and just push through the pain barrier when exercising. And it's a bit kind of counter intuitive, because when you are hurting and you don't really want to use that arm or leg or neck. But you have to really really push through it.” (P1) |
| 3.3 Family and friends offering support to get out of the house (shopping, movies etc)/ with ADLS | P1, P9, P5 | “…family and friends coming and visiting and trying to find things, you know, just even to, you know, take into the cinema just to make sure I go out and don’t just sit at home.” (P1)  “I mean, I've washed myself right the way down alone and then my wife just washes my lower leg and feet for me” (P5) |
| 3.4 Feeling grateful for all the input and support available to help get better | P1, P15, P5 x2 | “I'm just trying to be very positive, and I get a lot of encouragement, you know, from you and you know, from my physiotherapist. And I think this was just really amazing, you know, how many people you have around you just to try, you know, just to help you to get to get better” (P1)  “I think it were brilliant they got me up quite quickly yeah they were just they were just so reassuring they just make you feel like you’re safe” (P5)  “I am thankful for their help they put me on the right course and gave me that little bit of confidence which is you know what you really need. So yeah you know I’m really pleased with the way things are going” (P5) |
| 3.5 Received tailored care from community rehabilitation services to meet specific needs | P14, P1, P12 | “Oh look here's somebody who does sport I'm going to tailor what to say to him” (P14)  “Felt like being a complex case meant that people remember you and make an effort to help” (P1) |
| 3.6 Importance of getting good care in the community and the impact that has on your recovery | P14, P15, P7, P7, P12, P12 | “So all these things have just been happening successfully. Really great and they're all the things I wasn't expecting. So it's great news” (P14)  "The physio since being home is 10 times better than what I was getting in there.” (P7)  “So I phoned up the surgery, and they were very good, and gave me face to face that day. And the doctor gave me a little bit of feedback on you know her, take on what she saw.” (P12) |
| 3.7 How supported you feel by your health care professional depends on the individual - how invested and how competent they are. | P1 | “Yeah, I think it depends obviously on the person you know as anything. It's just. It's just who you know. Who you meet, and how invested they are, and how competent they are, how willing they are to help” (P1) |
| 3.8 Feel that earlier therapies interventions would have been beneficial | P2 | “Perhaps if I’d had these exercises a bit earlier on, instead of just now” (P2) |
| **Theme 4: Acknowledging the impact that critical illness has during admission, discharge and recovery on mental health of patient and family.** | | |
| 4.1 Acknowledging the mental trauma of what happened | P1, P12 | “To be honest, I don't want to go back there. You know. Be like. It makes me so anxious just thinking about it.” (P1)  “The mental support that I know I clearly need” (P12) |
| 4.2 No energy for doing anything but basic self care | P1 | ”Need weeks to build….[the] mental space in your brain to start doing other things” (P1) |
| 4.3 Family requiring support as part of the recovery process | P13 | “She witnessed the whole thing, and I'm more concerned about her… my daughter's responding quite good to the sessions.” (P13) |
| 4.4 Acknowledging the psychological role and determination in recovery | P5 | “Although it's a physical thing, it's also a mental thing, you know the recovery is also in your head you know if you feel safe in yourself than it doesn’t matter what other people say.” (P5)  “You know I don’t want to be walking around on these crutches blah blah blah. And they went well if anyone can do it you can we know that. That's you know, that's how people know me. Always been sort of that way you know determined” (P5) |
| 4.5 Believe I was discharged home too early from symptoms management perspective | P2 | “I think I was sent home a little bit too early… and that nearly sent me through the roof the pain. They thought, oh, good, she's walking. And obviously, you know, beds are needed for people who perhaps can't walk, and I could walk. So I was sent home” (P2) |
| 4.6 Discharge with family led to falling out | P2 | “It nearly caused the disruption in the family… I don't think he realised what he was taking on…I was left alone in a little room downstairs, and they were mostly being upstairs. I just felt, I just felt really, really terrible. I really thought I was going to die in my own son's house” (P2) |
| 4.7 Initially not feeling safe when discharged home - feeling of it being too early/ to the wrong place | P13 | “I didn't feel secure, but at the end, I mean, I can see that whatever I said, it didn't really make any difference. So it was time to get out of there and come home and not be somewhere where I wasn't really, really wanted after my discharge” (P13) |
| **Theme 5: Frustrations with fragmented care (including patient feeling not in control)** | | |
| 5.1 Care is fragmented and no-one is in overall control of my care | P9 x4, P2, P13, P6 | “For me, it's been a really exhausting part of recovery, that, that sense that you're on your own, and you've got to fight for a lot of things. And so that's, that's why and then I guess there's something on has the has the once you've left hospital, and you know, and community care, either by your GP or via other services? Are those services being delivered? Because there doesn't seem to me to be anyone in overall charge of things. It's quite fragmented. And you don't get the sense that anybody's in overall control, which is me, me, I'm the one in overall control, allegedly” (P9)  “There's quite there's, there's a lot of opportunities for people to fall through the cracks” (P9)  “Who would know if I was dead on the kitchen floor.” (P9)  “The biggest thing that's been a problem, not having a single point, point of contact” (P9)  “because I got so many issues right know, it's kind of hard to keep track of everything you're feeling” (P13) |
| 5.2 Fragmented care between providers | P7, P2 | “Went from the Royal London to another hospital and then a rehab unit and then home. It wasn’t just one transition” (P7)  “They did MRIs and everything, but they couldn't, they couldn't do any reports on it, because they didn't have anything to compare with…I don't think the two hospitals were able to talk to each other” (P2) |
| 5.3 Difficulties managing multiple follow-up appointments | P7 | “I can't remember when she said the appointment is. I need to check. Because so many appointments going on and I've tried to write a list down but you still miss stuff.” (P7) |
| 5.4 In early recovery, difficulty processing information about care planning | P7 | “You’re not always in a fit state to keep track of where it's all gone … The earlier on the worse you was so for a good half of that I wasn’t even fit to realise what was going on” (P7) |
| 5.5 Frustrations (and functional limitations) when not getting support services you need and therefore impacted on recovery | P9 (x9), P11, P7, P13 x2, P6 x2 | “I didn't know how long and how difficult it was going to get with the wrong stuff on the wheelchair. It just been the biggest issue.” (P9)  “And this is what was really frustrating with the community, they are hopeless and couldn't understand why I would ever want to leave my flat and start to use a toilet” (P9)  “This is a very expensive commode and you’ve got it and you need to use that. They couldn't understand why I would ever want to not use it” (P9)  “You can see where it's really creaking where the staffing levels just haven't been put into place” (P9)  “Not always very useful - I thought I could have figured that out myself.” (P9)  “My recovery, I mean, has been hampered by me not getting to see, I mean, the specialist I needed to see” (P13)  “You don't know the circumstances that make the person late. And then just said you are not seeing the person. If it is something that is more, they may have had casualty in their hands. You know, so…the GP surgery, they are not helping” (P6) |
| 5.6 Frustrations with new complications/symptoms since leaving hospital | P9, P15, P13, P6, P12x2 | “And then, of course, once the bump on my shoulder appeared, I mean, I wasn't so happy to use it anymore, you see, so but it's a kind of a, kind of a complicated situation” (P13)  “The doctor did not even ask me why did I come to the surgery? He just said…the blood the blood test you did is wrong because you have sickle cell, that is not the blood test to do. You have to come to the surgery, collect a blood something so that you can go again and he stopped, not even asking me any question. Or ask me if I have any question for him” (P6) |
| 5.7 No routine GP follow up or response to discharge papers. | P9, P15, P11, P8 | “And there wasn't any sense that anybody had proactively got my discharge report read it and thought, oh, gosh, we should probably contact this person.” (P9) |
| 5.8 Independently sourcing other services outside of the NHS to support to make up for lack of specialist input in community services | P9 x3, P7, P11, P2, P13 | “So I'm going to do it myself“ (P9)  “I’m fortunate in that I can afford just to do the stuff myself. I've I've got a good job. I can just pay for it. Not every most people aren't in that position. So it's frustrating to think there's people out there who are stuck in who are stuck unable to travel because yeah, that's seen as a luxury” (P9) |
| 5.9 Disengaged with services/ treatment as fed up | P7, P13 | “No, I discharged myself from the rehab, I’d had enough of being in there” (P7)  “I've got to take them for a valid reason that they're going to help me with the pain, which they're not so I prefer to disengage from them” (P13) |
| 5.10 Anxiety/ unease about therapies ending | P7 | “Cos the NHS physio is only 21 days. Unless they extend it which isn’t very long seeing my injuries” (P7) |
| 5.11 Declining therapy input due to lack of privacy | P13 | “I don't really want it to be quite honest. I don't want nobody, you know. I don't want nobody nosing around, as simple as that” (P13) |

**File 4: Clinician Codebook**

| **CODE** | Cited in | **QUOTES** |
| --- | --- | --- |
| **Recovery Process** | | |
| **Theme 1: Heterogenous nature of recovery from critical illness – with variations based upon admitting condition, patient and timescale factors** | | |
| 1.1 Wildly different focuses on recovery between when patient is discharged from ACCU to when they are seen in clinic | C2, C1 | “Yeah, wildly, I would say actually… they have no real idea of what their goals are and where they're going to end up when they leave here. And then there's this huge space of time in between them, leaving the doors of the fourth floor, going to the ward, going to rehab and going home before we see them again” (C2)  “Definitely critical care. You're very much the acute side of things, basically keeping somebody alive when you're doing the follow up clinic, it's much more rehab, psychological support, referrals that have been for, that have not been done, or that have gone missing, things that you wouldn't really deal with so much in the intensive care setting, I think.” (C1) |
| 1.2 Variety of different recovery pathways and timescales | C3, C5 | “Being an elective post-surgical unit majority of patients follow their expected pathway. Majority of them are awake. not delirious, progressing as anticipated along their elective recovery pathway. They stay with us for sort of between one and five days And then move out to the surgical ward and go home. Obviously there's because they're a high risk population, there's a reasonable amount of post-operative confusion, post-operative delirium, which might delay their path, but actually it's other post-operative complications that normally delay their recovery more” (C3) |
| 1.3 Complications may slow down recovery | C3 | “Post-op complications, you know, such as infections or ileus, et cetera, that might slow down that ITU stay critical care stay” (C3) |
| 1.4 Change from focus on physical recovery in hospital to psychological/ social in follow-up clinic | C3, C1 | “I imagine that their first thoughts it feels like their first thoughts are the physical but actually the sort of cognitive and psychological seem to make up as much, if not more of the stuff that we talk about [in follow-up clinic]” (C3) |
| 1.5 Sometimes ACCU clinicians are unsure how much someone will recover | C2 | “that's sometimes because we don't know, I guess, as well, how much you're going to progress” (C2) |
| 1.6 Potential for mismanagement of care once discharged from ACCU | C2 | “We pick up the pieces 12 months later and realize it's a bit of a disaster, and they've kind of dropped off from all other services” (C2) |
| 1.7 Potential for patients that fall through the cracks and be missed by services unless they seek out help themselves | C5 | “People kind of fall between the cracks haven't been picked up by services. And you know they're struggling much more to get on with things... They're not able to really Seek help unless they go into one of our services, and someone picks them up” (C5) |
| **Theme 2: The newness of recovering from critical illness limits patients’ understanding of the recovery process** | | |
| 2.1 Often patients don’t have a clear recovery trajectory at point of discharge from ACCU and hospital | C2, C1, C4 | “They have no real idea of what their goals are and where they're going to end up when they leave here” (C2)  “So most of them I think don't realize that basically I think that sometimes patients and their families think that I've left intensive care, I've left the hospital, I've had my rehab, and now I'm going home and it's going to be fine. But this is where most of the times the problem starts. And when I'm talking about problems and issues I mean fatigue and nightmares and anxiety and You know I think it's very difficult because it's the first time that they're kind of being alone without any healthcare professionals around them. It's just them and their family and friends. And I think that they don't realize up to that point that they do still have a long road ahead of them. And lots of challenges to face” (C4) |
| 2.2 Not many patients really understand how long their recovery can take | C2 | “Yeah, I think not many patients really understand how long it can take” (C2) |
| 2.3 Often patients don’t remember much about their stay on ACCU | C1, C5, C6 | “And then once they are awake, I'd say, speaking to them months down the line, often, they don't really remember much about their stay in intensive care” (C1)  “That often our patients might not have huge amounts of memory of their ICU admission.” (C6) |
| 2.4 Patients' post-op expectations of recovery process different to clinician | C3 | “Other times the patients are quite surprised. Like when you want to get them out of bed post-op day one after a major laparotomy, they're like, what? I've just had major surgery.” (C3)  “I think patients’ expectations before elective surgery are not as clear as we might have thought they were.” (C3) |
| 2.5 Realisation later on in recovery pathway that although physically functioning well, they may be struggling with other aspects (e.g. MH) | C5 | “Maybe I'll be able to manage this. I just want to get on with my life. They start going back to work. They start, you know, doing all the usual kind of life events and hobbies, etc. And then they start noticing that there are things that there aren't they aren't able to do.” (C5)  “People who've just tried to get on with everything, but actually, they're really struggling along and they could really do with some psychological help and support.” (C5) |
| 2.6 Not many patients have a clear understanding for potential of recovery | C2 x 3 | “Maybe don't really understand where they might reach” (C2)  “I think people don't necessarily understand always that it is a life changing injury that's not You're not going to go back to your baseline” (C2)  “They don't really understand where they might end up and what they might need to do to get a bit better” (C2) |
| 2.7 Often family have more of an awareness of trajectory/ recovery process so far than sick relative at point of discharge from ACCU  Family often are the ones to have discussions with clinical team about progress/recovery/trajectory as patient is critically unwell. | C1, C6, C3 | “They're definitely not as aware as then their relatives are more aware of kind of what priority was at that point, and the relative the patient doesn't necessarily realize how sick they were because they couldn't see it” (C1)  “But a lot of our interactions on the delirium ward round are actually more with the relatives than with the patient. Depends on the patient, but it is often with the relative” (C3) |
| 2.8 When is the right time to have discussion with patient about recovery process | C2 | “But then I also wonder about, when is the right time to get that message across?...I think it’s very patient specific” (C2)  “But I think it is difficult because some of our patients aren't able to have those conversations when they leave” (C2)  “I think that's not necessarily, because they {conversations about recovery} haven't happened, but because people aren't in the right place to receive that message whenever it was delivered” (C2) |
| 2.9 Uncertainty as to whether discussion about recovery process happens once left ACCU | C2 | “I think you go to the ward where there's a lot less input, and then the reality hits, maybe without anyone actually explaining to you what's gone on and that numbness in your legs probably never come back, or that, movement is never going to come back.” (C2) |
| 2.10 Clinicians aren’t always very good at communicating that patient will not return to their baseline | C2 | “I think we are probably not very good at perhaps getting that message across” (C2) |
| **Theme 3: Impact on, and lack of support for families** | | |
| 3.1 Lack of support (nationally) for family/relatives | C6 | “And I think sometimes we neglect it a little bit as well kind of ask families how they're doing, and. I think sometimes we do, but then we get a bit stuck, because if they then say. Actually, I'm not doing so well, I think about it a lot I haven't used it for. So I have nightmares. Then we're a bit like Hmm. Okay, what can we do with you now? Where can we. Okay? Because I think nationally, that's something that's lacking, anyway, is support. And families of people who have had severe illness critically. Illness” (C6)  “For loved ones, and I just don't have that capacity for it” (C5) |
| 3.2 Importance of recognising effect on family members (is asked in PICUPS) | C6, C5 | “Something I have been thinking about more recently is about support for families. And that's kind of related to the fact that often our patients might not have. Huge amounts of memory of their ICU admission. But their families do, because they were there. There they were awake throughout the whole thing. They were the ones who were. Told that their loved one might not survive or. There was, you know, seriously ill that they were, gonna have amputation, or whatever it was.” (C6)  “I'm fine, you know. I was the patient. I don't really remember or. You know. Actually, I'm getting on okay. But it's my. It's my partner or my son or my nephew that found me unconscious. And they're really struggling” (C5) |

**File 5: COREQ Checklist**

**Consolidated criteria for reporting qualitative studies (COREQ): 32-item checklist**

Developed from:

Tong A, Sainsbury P, Craig J. Consolidated criteria for reporting qualitative research (COREQ): a 32-item checklist for interviews and focus groups. International Journal for Quality in Health Care. 2007. Volume 19, Number 6: pp. 349 – 357

| **Item No** | **Guide Questions/Description** | **Reported on Page #** |  |  |
| --- | --- | --- | --- | --- |
| **Domain 1: Research team and reflexivity** | | |  |  |
| **Personal Characteristics** | | |  |  |
| 1. Interviewer/ facilitator | Which author/s conducted the interview or focus group? | Supplementary material, page 1 |  |  |
| 2. Credentials | What were the researcher’s credentials? E.g., PhD, MD | Supplementary material, page 1 |  |  |
| 3. Occupation | What was their occupation at the time of the study? | Supplementary material, page 1 |  |  |
| 4. Gender | Was the researcher male or female? | Supplementary material, page 1 |  |  |
| 5. Experience and training | What experience or training did the researcher have? | Supplementary material, page 1 |  |  |
| **Relationship with participants** | | |  |  |
| 6. Relationship established | Was a relationship established prior to study commencement? | Page 4 |  |  |
| 7. Participant knowledge of the interviewer | What did the participants know about the researcher? e.g. personal goals, reasons for doing the research? | Page 4 |  |  |
| 8. Interviewer characteristics | What characteristics were reported about the interviewer/facilitator? e.g. Bias, assumptions, reasons and interests in the research topic | Supplementary material, page 1 |  |  |
| **Domain 2: study design** | | |  |  |
| **Theoretical framework** | | |  |  |
| 9. Methodological orientation and Theory | What methodological orientation was stated to underpin the study? e.g. grounded theory, discourse analysis, ethnography, phenomenology, content analysis | Page 3 |  |  |
| **Participant selection** | | |  |  |
| 10. Sampling | How were participants selected? e.g., purposive, convenience, consecutive, snowball | Page 4 |  |  |
| 11. Method of approach | How were participants approached? e.g., face-to-face, telephone, mail, email | Page 4 |  |  |
| 12. Sample size | How many participants were in the study? | Page 4 |  |  |
| 13. Non-participation Setting | How many people refused to participate or dropped out? Reasons? | Page 4 |  |  |
| 14. Setting of data collection | Where was the data collected? e.g., home, clinic, workplace | Page 4 |  |  |
| 15. Presence of non-participants | Was anyone else present besides the participants and researchers? | Supplementary material, page 1 |  |  |
| 16. Description of sample | What are the important characteristics of the sample? e.g. demographic data, date | Page 4/5 |  |  |
| **Data collection** | | |  | No |
| 17. Interview guide | Were questions, prompts, and guides provided by the authors? Was it pilot tested? | Supplementary material, page 1 |  |  |
| 18. Repeat interviews | Were repeat interviews carried out? If yes, how many? | No |  |  |
| 19. Audio/visual recording | Did the research use audio or visual recording to collect the data? | Page 4 |  |  |
| 20. Field notes | Were field notes made during and/or after the interview or focus group? | No |  |  |
| 21. Duration | What was the duration of the interviews or focus group? | Page 4 |  |  |
| 22. Data saturation | Was data saturation discussed? | Page 4 |  |  |
| 23. Transcripts returned | Were transcripts returned to participants for comment and/or correction? | No |  |  |
| **Domain 3: analysis and findings** | | |  |  |
| **Data analysis** | | |  |  |
| 24. Number of data coders | How many data coders coded the data? | Page 4 |  |  |
| 25. Description of the coding tree | Did the authors provide a description of the coding tree? | Pages 6-7, 9-10 and full code-book in supplementary material, pages 3-20 |  |  |
| 26. Derivation of themes | Were themes identified in advance or derived from the data? | Page 4 |  |  |
| 27. Software | What software, if applicable, was used to manage the data? | None |  |  |
| 28. Participant checking | Did participants provide feedback on the findings? | No |  |  |
| **Reporting** | | |  |  |
| 29. Quotations presented | Were participant quotations presented to illustrate the themes/findings? Was each quotation identified? e.g., participant number | Pages 5-7, 8-10 and full code-book in supplementary material, pages 3-20 |  |  |
| 30. Data and findings consistent | Was there consistency between the data presented and the findings? | Pages 5-11 |  |  |
| 31. Clarity of major themes | Were major themes clearly presented in the findings? | Pages 5-11 |  |  |
| 32. Clarity of minor themes | Is there a description of diverse cases or a discussion of minor themes? | Pages 5-11 |  |  |
